# Diagnostic accuracy of Faecal Immunochemical Testing for patients with symptoms of colorectal cancer: a retrospective cohort study of 14,487 consecutive test requests from English primary care

**DOI:** 10.1101/2020.05.15.20077909

**Authors:** Brian D Nicholson, Tim James, Maria Paddon, Steve Justice, Jason L Oke, James E East, Brian Shine

## Abstract

**Objective:** To ascertain the diagnostic performance of faecal immunochemical test (FIT) in symptomatic primary care patients, to provide objective data on which to base referral guidelines.

**Design:** Stool samples from routine primary care practice in Oxfordshire, UK were analysed using the HM-JACKarc FIT method between March 2017 to March 2020. Clinical details described on the test request were recorded. Patients were followed up for up-to 36 months in linked hospital records for evidence of benign and serious (colorectal cancer, high-risk adenomas and bowel inflammation) colorectal disease. The diagnostic accuracy of FIT is reported by gender, age, and FIT threshold.

**Results:** In 9,896 adult patients with at least 6 months of follow-up, a FIT result ≥10 μg/g had an overall sensitivity for colorectal cancer of 90.5% (95% CI 84.9%-96.1%), women 90.0 (80.7-99.3), men 90.8 (83.7-97.8); overall specificity 91.3 (90.8-91.9), women 92.4 (91.8-93.1), men 89.8 (88.8-90.7); overall Positive Predictive Value (PPV) 10.1 (8.15-12.0), women 7.64 (5.24-10.0), men 12.5 (9.52-15.5)); and an overall Negative Predictive Value (NPV) 99.9 (99.8-100.0), women 99.8 (99.7-100.), men 99.9 (99.9-100.0). The PPV and specificity of FIT were higher for serious colorectal disease combined and the sensitivity and NPV were lower than for colorectal cancer alone. The Area Under the Curve (AUC) for all patients did not change substantially by increasing the minimum age of testing. In this population, 10% would be further investigated to detect 91% of the cancers at 10ug/g and 3% further investigated to detect 54% of the cancers at 150ug/g. The number needed to scope to detect one cancer was ten using FIT at 10ug/g.

**Conclusion:** A FIT threshold of 10 µg/g is appropriate to triage adult patients presenting to primary care with symptoms of serious colorectal disease. FIT may provide an appropriate approach to reprioritising patients colorectal cancer symptoms whose tests have been delayed by the COVID-19 pandemic.

**What is already known on this subject?:** Faecal Immunochemical Testing (FIT) is recommended by NICE to triage symptomatic primary care patients into further investigation for serious colorectal disease, including colorectal cancer. Almost no real-world data exists documenting the diagnostic accuracy of low FIT thresholds associated with colorectal cancer or serious colorectal disease in primary care with symptoms of colorectal cancer.

**What are the new findings?:** In 9,896 consecutive FITs submitted by English General Practitioners to a large English laboratory, using a threshold of 10ug/g, FIT had a sensitivity and specificity of 91% for colorectal cancer, a sensitivity of 53% and a specificity of 92% for serious colorectal disease.

Of the population tested with FIT, 10% would be further investigated to detect 91% of the cancers at 10ug/g, 4% would be further investigated to detect 74% of the cancers at 50ug/g, and 3% further investigated to detect 54% of the cancers at 150ug/g.

**How might it impact on clinical practice in the foreseeable future?:** Low threshold FIT could be used as a triage test without overburdening endoscopy resources, supporting widespread implementation of the NICE recommendations for its use in low-risk patients in primary care.

FIT may be an effective test for re-prioritizing patients whose endoscopy test have been deferred due to the COVID-19 pandemic to match available endoscopy resources to those at highest risk of colorectal cancer.

## Introduction

Since July 2017 the Faecal Immunochemical Test (FIT) has been recommended by the UK’s NICE DG30 guidelines “to guide referral for suspected colorectal cancer in people without rectal bleeding who have unexplained symptoms but do not meet the criteria for a suspected cancer pathway” (1). The adoption of FIT in primary care has been slow, with notable variation in uptake and implementation across the UK (4). Guidance on faecal testing had generated significant debate (2, 3). Concerns have been raised about delayed cancer diagnosis due to false negatives and the potential to increase demand on already stretched endoscopy service due to false positives, a common problem associated with the previously used guaiac (gFOB) method (4). FIT is more specific than gFOB as it is an immunoassay based method that measures the globin component of human haemoglobin and its early degradation products (5, 6). This means it does not require dietary restriction, is specific to lower gastrointestinal (GI) cancers as upper GI enzymes degrade human globin, and is less affected by concomitant medication use. Accordingly, three times fewer false positive tests are reported when FIT is compared to gFOB in samples sent to the laboratory for symptomatic patients (7).

NICE recommended a quantitative FIT threshold of 10 µg/g faeces should trigger referral for colonoscopy (1). The UK Bowel Cancer Screening Programme (UKBCSP) currently uses a threshold of 120 µg/g faeces. Colorectal cancer screening studies have consistently demonstrated improved performance of FIT compared to gFOB at a high analytical threshold (8-10). However, NICE expressed concerns about the applicability of all ten of the studies used to underpin their DG30 recommendation: none reported data on people with low risk symptoms of colorectal cancer (1) and only one study was conducted in primary care (11).

There has since been evidence published in support the use of FIT at a lower threshold in symptomatic primary care patients. A small English comparative study showed similar sensitivity and improved specificity for FIT for colorectal cancer compared to gFOB in primary care patients with low-risk symptoms (7). A Danish study concluded that FIT may be used as a supplementary diagnostic test in individuals with non-alarm symptoms of colorectal cancer to diagnose serious bowel disease (colorectal cancer, high risk adenoma, and inflammatory bowel disease) (12). Authors of a Scottish study strongly recommended FIT should become integral to the assessment of all patients presenting to primary care with new bowel symptoms, to objectively determine the risk of underlying serious bowel disease (13). Despite these important studies, data from symptomatic primary care patients prior to referral is lacking. Unanswered questions remain: does FIT perform similarly in men and women, across age-groups, and what is the optimal FIT threshold to detect colorectal cancer and significant gastrointestinal disease? (14,15).

The COVID-19 pandemic has introduced a new urgency to identify non-invasive approaches to triage for patients with symptoms of serious colorectal disease requiring further investigation. The large backlog of endoscopy created by the COVID-19 pandemic means it is almost certain the widespread use of low cut-off FIT testing will be required to risk stratify all patients referred with possible cancer symptoms into groups for urgent and less urgent endoscopy immediately after the pandemic has passed. Understanding the performance of FIT at a range of thresholds in symptomatic patients is therefore a priority.

The Oxford University Hospitals Trust (OUH) first adopted FIT prior to the DG30 guidance to comply with the 2015 NG12 NICE guidance for suspected cancer (6). This coincided with a desire from the clinical laboratory to move away from gFOB. FIT was commissioned by Oxfordshire Clinical Commissioning Group (OCCG) as a direct access test for General Practitioners in 2016 following a study by this group comparing the accuracy of the gFOB and FIT methods in symptomatic primary care patients meeting NG12 criteria (5). We conducted a diagnostic accuracy study using linked electronic hospital records data to establish the utility of FIT to detect serious bowel disease in the context of the NICE DG30 guidelines in England. We considered the utility of FIT by presenting symptom, age-group and gender, to identify the optimal FIT threshold, and documented FIT negative cases of colorectal cancer.

## Methods

This retrospective study included consecutive FIT samples sent to the laboratory from primary care in the period March 2017 to March 2020. The Oxford University Hospitals NHS Foundation Trust (OUHT) Clinical Biochemistry laboratory serves all primary care clinicians in the county of Oxfordshire with a population of approximately 660,000. The department is one of the largest in the United Kingdom, performing over 8 million tests a year, and has an ‘M’-based laboratory information management system. All FIT testing is undertaken in a single laboratory at the John Radcliffe Hospital. The assessment was registered as a service evaluation on our hospitals Datix register (CSS-BIO-3 4730). We followed STARD reporting guidelines (16).

Leading up to the study period, the change in NICE guidance and the indications for FOB testing were communicated to GPs in Oxfordshire by email and newsletter from the Oxfordshire Clinical Commissioning Group (OCCG). Samples were collected into standard stool pots by patients in primary care and analysed for FIT using the HM-JACKarc analyser (Kyowa Medex Co., Ltd., Tokyo, Japan) a method that has been independently evaluated with respect to analytical performance (16) and is recommended in the context of use for samples from primary care (1). The method had a calibration range of 7 to 450 µg Hb/g faeces and immunoassay reproducibility, assessed across 12 months was between 4.5% and 8.7% when expressed as a percentage coefficient of variation. Sample preparation prior to analysis on the FIT instrument utilized the Extel Hemo-Auto MC device, a process which introduced additional variation, with overall analytical imprecision observed to be between 7.0 and 13.5% when specimens had been homogenised and sampled by laboratory staff. The selection of the Hb concentration considered positive was made before the NICE recommendation to use 10 ng/g and was based on the methods lowest calibrant value of 7 µg/g and agreed with the OCCG based on initial method verification data (7). Results were reported electronically to the requesting GP as either positive or negative. In selecting the approach to faecal sample handling we balanced two competing pre-analytical sources of error: the requirements to minimize sampling errors if undertaken by the patient, which may give rise to false negatives if the collection device was inadequately filled; and specimen degradation concerns if sampling is undertaken in the laboratory due to delays between specimen collection and stabilisation in the collection device buffer. We have highlighted the balancing of these risks in our contribution to the NICE FIT adoption resource (1). Where more than one sample result was available for any individual patient, any positive result within those samples tested was considered a positive outcome on the basis that a single positive would trigger referral.

To confirm the presence or absence of disease, OUHT clinical and diagnostic databases were searched for evidence of cellular pathology for up to 36 months following the FIT test for all patients. Histology, endoscopy and CT colonoscopy reports were retrieved by searching by both hospital and NHS number. Patients were classified individually then by discussion between members of the research team (BS, BN, TJ, JE) having colorectal cancer, normal cellular pathology findings, colorectal polyps, inflammation of the colon, or no further follow-up investigation for between 6 and 36 months. Patients who had no further investigation were categorised as negative for serious pathology as any serious pathology would be expected to have presented to secondary care within this time period. Serious disease was a combination of any of: colorectal cancer, large polyp or high-grafe dysplasia, or inflammation of the colon.

The diagnostic accuracy of FIT in relation to a diagnosis of cancer within six months confirmed within the linked hospital record was summarised using sensitivity, specificity, negative and positive predictive values, the area under the curve (AUC) and exact 95% confidence intervals. A sensitivity analysis investigated the effect of using follow-up periods of three, six and 12 months. We used the sensitivity and specificity and prevalence of colorectal cancer from this analysis to derive estimates of positive and negative FIT tests in relation to cancer and serious colorectal disease outcomes per 1000 patients tested. In addition, the clinical details of patients with a negative FIT were collated.

## Results

14,487 consecutive FITs were conducted in 12,509 adults during the study period. 9,896 patients had at least six months of follow-up and were retained in the primary analysis. The median age was 60 years (range 18-101 years, inter-quartile range 51-74 years). 5,795 (58.6%) were women (59 (18-101) [51-74]) and 4,101 (41.4%) were men (62 (18-99) [52-76]). A larger proportion of FITs were positive (≥7µg/g) in men (13.4%) than women (9.6%), and in older people (e.g. 18.8% of women aged ≥80 years compared to 8.7% aged 18-39 years) (Table 1).

**Table 1:** Clinical details of people tested with FIT.

|  | **Women**<br>N (% FIT ≥7<br>µg/g) | **Men**<br>N (% FIT ≥7<br>µg/g) | **All**<br>N (% FIT ≥7<br>µg/g) |
| --- | --- | --- | --- |
| **Age-group (years)** |  |  |  |
| 18-39 | 459 (7.41) | 334 (10.8) | 793 (8.8) |
| 40-49 | 891 (6.06) | 509 (9) | 1400 (7.1) |
| 50-59 | 1681 (6.72) | 1068 (9.7) | 2749 (7.9) |
| 60-69 | 947 (7.39) | 704 (11.4) | 1651 (9.1) |
| 70-79 | 1047 (12.7) | 821 (16.1) | 1868 (14.2) |
| ≥80 | 770 (18.8) | 665 (22.7) | 1435 (20.6) |
| **Clinical Features** |  |  |  |
| Abdominal pain | 1,488 (6.99) | 1,013 (9.18) | 2,501 (7.88) |
| Anaemia | 1,587 (12.30) | 1,204 (17.40) | 2,791 (14.5) |
| Blood in stools | 805 (17.00) | 672 (22.90) | 1,477 (19.7) |
| Change in bowel habit | 3,000 (6.67) | 2,011 (8.95) | 5,011 (7.58) |
| Inflammation | 114 (17.50) | 59 (20.30) | 173 (18.5) |
| Iron deficiency | 772 (10.10) | 436 (14.40) | 1,208 (11.7) |
| Thrombocytosis | 99 (6.06) | 35 (14.3) | 134 (8.21) |
| Tired all the time | 33 (6.06) | 33 (9.09) | 66 (7.58) |
| Weight loss | 488 (9.63) | 463 (12.1) | 951 (10.8) |
| **Outcome** |  |  |  |
| Colorectal cancer | 40 (90%) | 65 (92.3%) | 105 (91.4%) |
| Benign disease (Polyp <10mm/Diverticulosis) | 40 (15%) | 40 (30%) | 80 (22.5%) |
| Bowel inflammation | 116 (27.6%) | 88 (47.7%) | 204 (36.3%) |
| High risk adenoma (Polyp >10mm or high-grade dysplasia) | 186 (40.3%) | 187 (42.8%) | 373 (41.6%) |
| No significant pathology | 255 (14.9%) | 163 (18.4%) | 418 (16.3%) |
| No further investigation | 5,158 (7.02%) | 3,558 (9.13%) | 8,716 (7.88%) |
| **Total** | **5795 (9.5)** | **4101 (13.4)** | **9896 (11.1)** |

### Clinical details and outcomes

FIT requests included a range of clinical features (symptoms, signs, test results) that gave rise to a concern about serious colorectal pathology. Patients commonly presented with combinations of clinical features (Table 1). The most common clinical features were change in bowel habit (included in 50.6% of requests), anaemia (28.2%), abdominal pain (25.2%), blood in stools (19.7%), and iron deficiency (12.2%). Significant colorectal disease was detected in 682 (6.9%) of patients tested, 105 (1.1%) were diagnosed with colorectal cancer, 373 (3.8%) large >10mm or high-grade dysplastic polyps, and 204 (2.1%) patients who had bowel inflammation (Table 1). Figure 1 shows the FIT values according to outcome group and gender.

**Figure 1.**
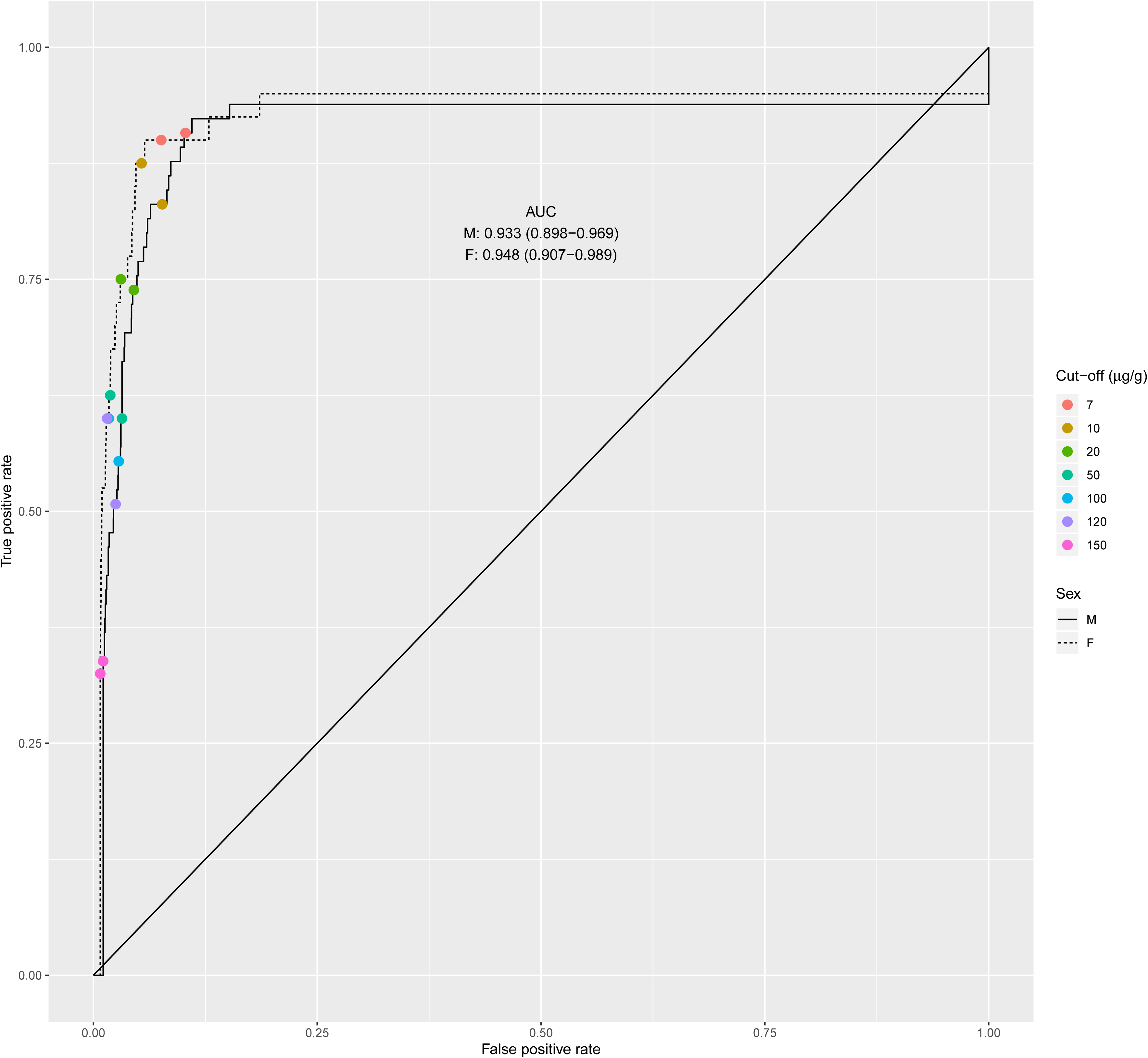
Quantitative FIT results by outcome category and gender. F: female; M: male.

### Overall diagnostic accuracy – gender and age

Table 2 presents AUCs by age cut-off for men and for women for both a diagnosis of colorectal cancer and for significant colorectal disease. Increasing the lower age-limit for FIT testing made very little overall difference to the overall AUC for colorectal cancer with poorer discrimination noted when using a higher cut-off. The AUC for colorectal cancer was 0.941 (0.914-0.968) ranging from 0.886 (0.805-0.967) in men aged ≥80 years, to 0.933 (0.898-0.969) in men aged ≥18yrs, and 0.881 (0.693-1) in women aged ≥80 years, to 0.948 (0.907-0.989) in women aged >18 years.

**Table 2:** Area Under the Curve (AUC) for FIT for a diagnosis of cancer or serious colorectal disease by gender and age-group.

| **Colorectal Cancer** |  |  |  |
| --- | --- | --- | --- |
| **Age-group** | **Women** | **Men** | **All** |
| ≥18yrs | 0.948 (0.907-0.989) | 0.933 (0.898-0.969) | 0.941 (0.914-0.968) |
| ≥40yrs | 0.944 (0.899-0.988) | 0.934 (0.897-0.972) | 0.940 (0.911-0.968) |
| ≥50yrs | 0.938 (0.889-0.987) | 0.929 (0.887-0.971) | 0.934 (0.903-0.965) |
| ≥60yrs | 0.919 (0.856-0.982) | 0.921 (0.870-0.971) | 0.921 (0.882-0.960) |
| ≥70yrs | 0.934 (0.867-1.000) | 0.936 (0.895-0.978) | 0.937 (0.901-0.972) |
| ≥80yrs | 0.881 (0.693-1.000) | 0.886 (0.805-0.967) | 0.889 (0.812-0.965) |
| **Serious colorectal disease** |  |  |  |
| ≥18yrs | 0.707 (0.663-0.751) | 0.781 (0.738-0.825) | 0.744 (0.713-0.775) |
| ≥40yrs | 0.698 (0.652-0.744) | 0.769 (0.723-0.816) | 0.733 (0.701-0.766) |
| ≥50yrs | 0.708 (0.658-0.758) | 0.769 (0.719-0.819) | 0.740 (0.704-0.775) |
| ≥60yrs | 0.648 (0.586-0.710) | 0.773 (0.712-0.833) | 0.711 (0.667-0.756) |
| ≥70yrs | 0.662 (0.581-0.743) | 0.793 (0.722-0.864) | 0.731 (0.676-0.786) |
| ≥80yrs | 0.649 (0.508-0.790) | 0.764 (0.655-0.874) | 0.716 (0.628-0.804) |

### Diagnostic accuracy by FIT threshold

For a colorectal cancer diagnosis in adults, the sensitivity of FIT decreased from 91.4 (86.1-96.8) at a cut-off of 7μ/g to 54.3 (44.8-63.8) at 150μ/g (Figure 2, Table 3). In women, the sensitivity decreased from 90.0 (80.7-99.3) at 7μ/g to 60.0 (44.8-75.2) at 150µg/g (Figure 2, Table 3). For men, sensitivity was 92.3 (85.8-98.8) at 7µg/g decreasing to 50.8 (38.6-62.9) at 150µg/g. Specificity in adults increased from 89.8 (89.2-90.4) at ^7^Hg/g to 98.1 (97.8-98.4) at 150µg/g, in women from 91.1 (90.3-91.8) to 98.5 (98.2-98.8), and in men from 87.9 (86.9-88.9) at 7µg/g to 97.5 (97.1-98.0) at 150µg/g in men.

**Figure 2:**
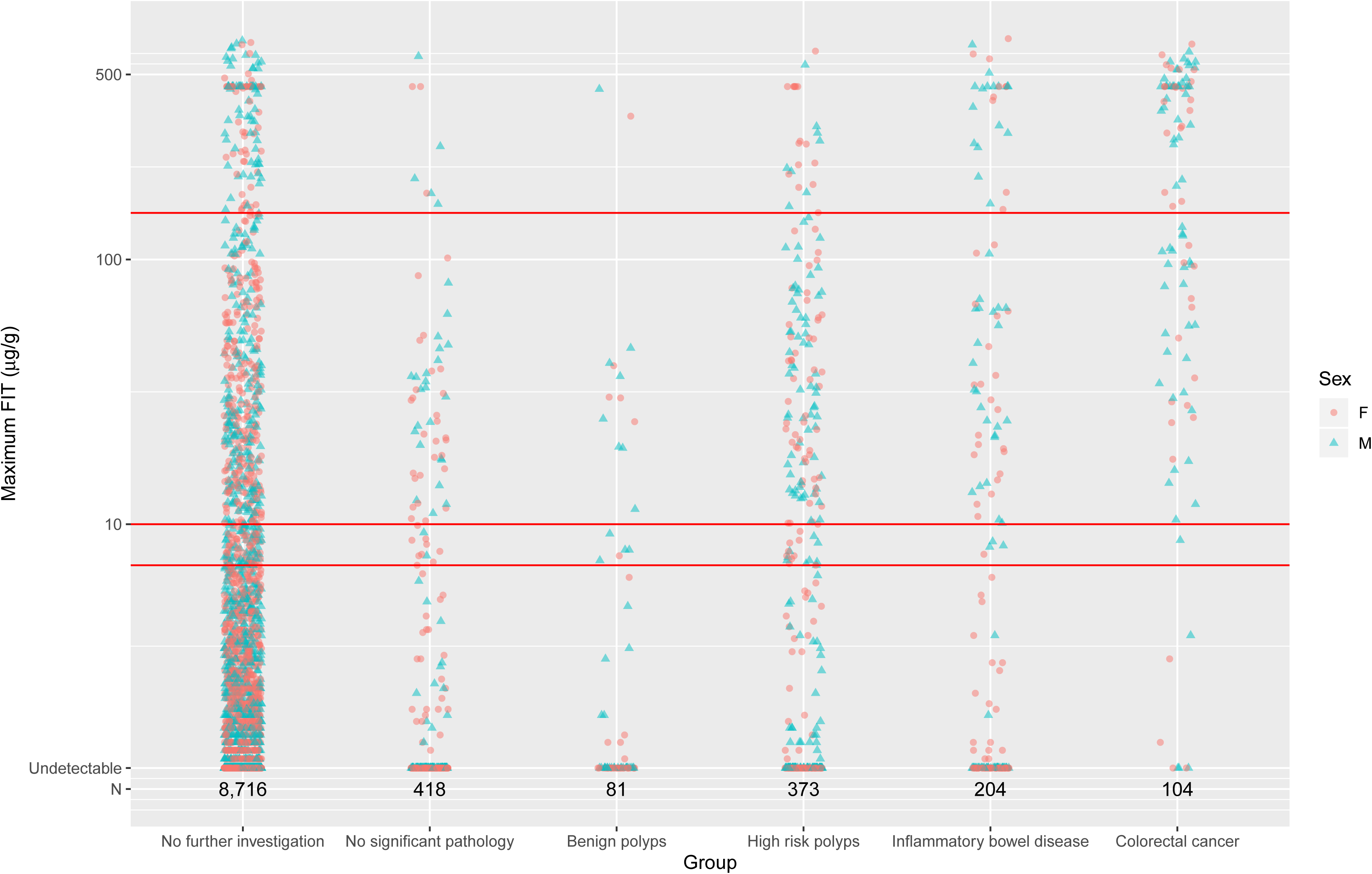
Receiver operating characteristic (ROC) curves of FIT for the detection of colorectal cancer. Coloured symbols correspond to the true positive / false positive rate estimates for thresholds of 7,10,20,50,100,120 and 150 μg/g. AUC = area under the curve.

In all adults, the PPV was 8.74 (7.07-10.4) at 7µg/g and 23.4 (18.1-28.7) at 150µg/g (Table 3). The PPV was greater in men than in women, especially at lower thresholds. In women, the PPV was 6.56 (4.49-8.63) at 7µg/g compared to 10.9 (8.32-13.5) in men. At 150µg/g the PPV was 21.4 (13.8-29.0) in women and 25.0 (17.6-32.4) in men. The NPV for colorectal cancer remained ≥99% for men and women at all thresholds studied (Table 3). Increasing the threshold from 7µg/g to 150µg/g reduced the NPV from 99.9 (99.8-100.0) to 99.5 (99.4-99.6) overall. This was similar for women (99.9 (99.8-100.) to 99.7 (99.6-99.9)) and men (99.9 (99.7-100.0) to 99.2 (98.9-99.5)).

**Table 3.** Comparison of diagnostic accuracy of FIT by cut-off for the detection of colorectal cancer. PPV: positive predictive value; NPV: negative predictive value.

| Colorectal Outcome | FIT cut-off (µg/g) | Women |  |  |  | Men |  |  |  | All |  |  |  |
| --- | --- | --- | --- | --- | --- | --- | --- | --- | --- | --- | --- | --- | --- |
|  |  | Sensitivity (95% CI) | Specificity (95% CI) | PPV (95% CI) | NPV (95% CI) | Sensitivity (95% CI) | Specificity (95% CI) | PPV (95% CI) | NPV (95% CI) | Sensitivity (95% CI) | Specificity (95% CI) | PPV (95% CI) | NPV (95% CI) |
| Cancer | 7 | 90.0<br>(80.7-99.3) | 91.1<br>(90.3-91.8) | 6.56<br>(4.49-8.63) | 99.9<br>(99.8-100.) | 92.3<br>(85.8-98.8) | 87.9<br>(86.9-88.9) | 10.9<br>(8.32-13.5) | 99.9<br>(99.7-100.) | 91.4<br>(86.1-96.8) | 89.8<br>(89.2-90.4) | 8.74<br>(7.07-10.4) | 99.9<br>(99.8-100.) |
|  | 10 | 90.0<br>(80.7-99.3) | 92.4<br>(91.8-93.1) | 7.64<br>(5.24-10.0) | 99.9<br>(99.9-100.) | 90.8<br>(83.7-97.8) | 89.8<br>(88.8-90.7) | 12.5<br>(9.52-15.5) | 99.8<br>(99.7-100.) | 90.5<br>(84.9-96.1) | 91.3<br>(90.8-91.9) | 10.1<br>(8.15-12.0) | 99.9<br>(99.8-100.) |
|  | 20 | 87.5<br>(77.3-97.7) | 94.6<br>(94.1-95.2) | 10.2<br>(7.00-13.4) | 99.9<br>(99.8-100.) | 83.1<br>(74.0-92.2) | 92.3<br>(91.5-93.2) | 14.9<br>(11.2-18.5) | 99.7<br>(99.5-99.9) | 84.8<br>(77.9-91.6) | 93.7<br>(93.2-94.2) | 12.6<br>(10.2-15.1) | 99.8<br>(99.7-99.9) |
|  | 50 | 75.0<br>(61.6-88.4) | 96.9<br>(96.5-97.4) | 14.6<br>(9.75-19.4) | 99.8<br>(99.7-99.9) | 73.8<br>(63.2-84.5) | 95.5<br>(94.9-96.2) | 21.0<br>(15.7-26.2) | 99.6<br>(99.4-99.8) | 74.3<br>(65.9-82.6) | 96.4<br>(96.0-96.7) | 17.9<br>(14.3-21.5) | 99.7<br>(99.6-99.8) |
|  | 100 | 62.5<br>(47.5-77.5) | 98.1<br>(97.8-98.5) | 18.9<br>(12.3-25.6) | 99.7<br>(99.6-99.9) | 60.0<br>(48.1-71.9) | 96.8<br>(96.3-97.3) | 23.2<br>(16.8-29.6) | 99.3<br>(99.1-99.6) | 61.0<br>(51.6-70.3) | 97.6<br>(97.3-97.9) | 21.3<br>(16.7-26.0) | 99.6<br>(99.4-99.7) |
|  | 120 | 60.0<br>(44.8-75.2) | 98.3<br>(98.0-98.6) | 19.8<br>(12.7-26.9) | 99.7<br>(99.6-99.9) | 55.4<br>(43.3-67.5) | 97.2<br>(96.7-97.7) | 24.0<br>(17.2-30.8) | 99.3<br>(99.0-99.5) | 57.1<br>(47.7-66.6) | 97.8<br>(97.6-98.1) | 22.1<br>(17.2-27.1) | 99.5<br>(99.4-99.7) |
|  | 150 | 60.0<br>(44.8-75.2) | 98.5<br>(98.2-98.8) | 21.4<br>(13.8-29.0) | 99.7<br>(99.6-99.9) | 50.8<br>(38.6-62.9) | 97.5<br>(97.1-98.0) | 25.0<br>(17.6-32.4) | 99.2<br>(98.9-99.5) | 54.3<br>(44.8-63.8) | 98.1<br>(97.8-98.4) | 23.4<br>(18.1-28.7) | 99.5<br>(99.4-99.6) |

For the serious colorectal disease grouping, sensitivity was lower than for colorectal cancer across all thresholds studied, ranging from 55.0 (49.5-60.6) at 7µg/g to 26.9 (21.9-31.8) at 150µg/g (Table 4). The NPV was also lower ranging from 98.4 (98.2-98.7) to 97.7 (97.4-98.0), whereas the specificity was higher 90.3 (89.7-90.9) to 98.3 (98.1-98.6), and the PPV was higher 15.5 (13.3-17.6) to 34.0 (28.1-40.0).

**Table 4.** Comparison of diagnostic accuracy of FIT by cut-off for the detection of serious colorectal disease (cancer; high risk adenoma; bowel inflammation). PPV: positive predictive value; NPV: negative predictive value.

| Colorectal Outcome | FIT cut-off (µg/g) | Women |  |  |  | Men |  |  |  | All |  |  |  |
| --- | --- | --- | --- | --- | --- | --- | --- | --- | --- | --- | --- | --- | --- |
|  |  | Sensitivity (95% CI) | Specificity (95% CI) | PPV (95% CI) | NPV (95% CI) | Sensitivity (95% CI) | Specificity (95% CI) | PPV (95% CI) | NPV (95% CI) | Sensitivity (95% CI) | Specificity (95% CI) | PPV (95% CI) | NPV (95% CI) |
| Serious disease | 7 | 43.6<br>(35.8-51.4) | 91.5<br>(90.7-92.2) | 12.4<br>(9.63-15.1) | 98.3<br>(98.0-98.7) | 66.7<br>(59.2-74.1) | 88.7<br>(87.7-89.7) | 18.6<br>(15.3-21.8) | 98.6<br>(98.2-99.0) | 55.0<br>(49.5-60.6) | 90.3<br>(89.7-90.9) | 15.5<br>(13.3-17.6) | 98.4<br>(98.2-98.7) |
|  | 10 | 42.9<br>(35.2-50.7) | 92.8<br>(92.2-93.5) | 14.2<br>(11.1-17.4) | 98.3<br>(98.0-98.7) | 64.1<br>(56.4-71.7) | 90.5<br>(89.6-91.4) | 20.8<br>(17.1-24.4) | 98.5<br>(98.1-98.9) | 53.4<br>(47.8-59.0) | 91.9<br>(91.3-92.4) | 17.5<br>(15.1-19.9) | 98.4<br>(98.1-98.7) |
|  | 20 | 36.5<br>(29.0-44.1) | 94.9<br>(94.4-95.5) | 16.6<br>(12.7-20.6) | 98.2<br>(97.8-98.5) | 57.5<br>(49.7-65.3) | 93.0<br>(92.2-93.8) | 24.2<br>(19.8-28.7) | 98.3<br>(97.8-98.7) | 46.9<br>(41.4-52.5) | 94.1<br>(93.7-94.6) | 20.5<br>(17.6-23.5) | 98.2<br>(97.9-98.5) |
|  | 50 | 28.8<br>(21.7-36.0) | 97.1<br>(96.7-97.6) | 21.8<br>(16.2-27.5) | 98.0<br>(97.6-98.4) | 47.1<br>(39.1-55.0) | 96.0<br>(95.4-96.6) | 31.4<br>(25.4-37.5) | 97.9<br>(97.5-98.4) | 37.9<br>(32.5-43.3) | 96.7<br>(96.3-97.0) | 26.9<br>(22.7-31.1) | 98.0<br>(97.7-98.3) |
|  | 100 | 23.7<br>(17.0-30.4) | 98.3<br>(98.0-98.7) | 28.0<br>(20.4-35.7) | 97.9<br>(97.5-98.3) | 36.6<br>(29.0-44.2) | 97.2<br>(96.6-97.7) | 33.3<br>(26.2-40.5) | 97.5<br>(97.0-98.0) | 30.1<br>(25.0-35.2) | 97.8<br>(97.5-98.1) | 31.0<br>(25.8-36.2) | 97.7<br>(97.5-98.0) |
|  | 120 | 21.8<br>(15.3-28.3) | 98.5<br>(98.1-98.8) | 28.1<br>(20.1-36.1) | 97.8<br>(97.5-98.2) | 34.0<br>(26.5-41.5) | 97.5<br>(97.0-98.0) | 34.7<br>(27.1-42.3) | 97.4<br>(97.0-97.9) | 27.8<br>(22.8-32.8) | 98.1<br>(97.8-98.3) | 31.7<br>(26.2-37.3) | 97.7<br>(97.4-98.0) |
|  | 150 | 21.8<br>(15.3-28.3) | 98.6<br>(98.3-98.9) | 30.4<br>(21.8-38.9) | 97.9<br>(97.5-98.2) | 32.0<br>(24.6-39.4) | 97.9<br>(97.5-98.3) | 37.1<br>(28.9-45.4) | 97.4<br>(96.9-97.9) | 26.9<br>(21.9-31.8) | 98.3<br>(98.1-98.6) | 34.0<br>(28.1-40.0) | 97.7<br>(97.4-98.0) |

### Trade-offs per 1000 patients tested

If all patients were referred and received definitive testing for colorectal cancer, using a FIT threshold of 10µg/g would lead to seven people without cancer referred for each person referred with cancer (Table 5). In women this ratio was twelve to one. In men, seven to one. Fewer people with a positive FIT of ≥10µg/g would need to be referred to detect one serious colorectal disease: two to one overall, increasing to three to one in women, and decreasing back to two to one in men (Table 5). However, although a lower number of patients would need to be investigated further to detect serious colorectal disease using a FIT threshold of ≥10µg/g, only 53% of the patients in the population with serious colorectal disease would be further investigated compared to 90% of people with colorectal cancer.

**Table 5:** Patients with cancer detected if patients with positive FIT referred expressed by FIT threshold and gender per 1000 patients FIT tested.

|  | **Threshold**<br>**(µg/g)** | **Positive**<br>**FITs**<br>**n (%)** | **Negative**<br>**FITs**<br>**n (%)** | **Cancers**<br>**detected**<br>**n (%)** | **Positive FITs**<br>**to detect one**<br>**cancer (number**<br>**needed to**<br>**scope)** | **Patients with cancer**<br>**and a negative FIT**<br>**(the cancer miss rate**<br>**per 1000 tests)** | **Serious disease**<br>**detected**<br>**n (%)** | **Positive FITs**<br>**to detect one**<br>**serious colorectal**<br>**disease (number**<br>**needed to scope)** |
| --- | --- | --- | --- | --- | --- | --- | --- | --- |
| Women | **7** | 95 (9) | 905 (91) | 6.2 (90) | 15 | 1 | 25.7 (44) | 4 |
|  | **10** | 82 (8) | 918 (92) | 6.2 (90) | 13 | 1 | 25.3 (43) | 4 |
|  | **20** | 60 (6) | 940 (94) | 6 (88) | 10 | 1 | 21.5 (37) | 3 |
|  | **50** | 36 (4) | 964 (96) | 5.2 (75) | 7 | 2 | 17 (29) | 3 |
|  | **100** | 23 (2) | 977 (98) | 4.3 (63) | 5 | 3 | 14 (24) | 2 |
|  | **120** | 21 (2) | 979 (98) | 4.1 (60) | 5 | 3 | 12.9 (22) | 2 |
|  | **150** | 19 (2) | 981 (98) | 4.1 (60) | 5 | 3 | 12.9 (22) | 2 |
| Men | **7** | 134 (13) | 866 (87) | 14.6 (92) | 9 | 1 | 55.3 (67) | 3 |
|  | **10** | 115 (11) | 885 (89) | 14.4 (91) | 8 | 2 | 53.1 (64) | 3 |
|  | **20** | 89 (9) | 911 (91) | 13.2 (83) | 7 | 3 | 47.7 (58) | 2 |
|  | **50** | 56 (6) | 944 (94) | 11.7 (74) | 5 | 4 | 39 (47) | 2 |
|  | **100** | 41 (4) | 959 (96) | 9.5 (60) | 4 | 7 | 30.3 (37) | 2 |
|  | **120** | 36 (4) | 964 (96) | 8.8 (55) | 4 | 7 | 28.2 (34) | 2 |
|  | **150** | 33 (3) | 967 (97) | 8.1 (51) | 4 | 8 | 42.1 (51) | 1 |
| All | **7** | 111 (11) | 889 (89) | 9.7 (91) | 11 | 1 | 37.9 (55) | 3 |
|  | **10** | 96 (10) | 904 (90) | 9.6 (91) | 10 | 1 | 36.8 (53) | 3 |
|  | **20** | 71 (7) | 929 (93) | 9 (85) | 8 | 2 | 32.3 (47) | 3 |
|  | **50** | 44 (4) | 956 (96) | 7.9 (74) | 6 | 3 | 26.1 (38) | 2 |
|  | **100** | 30 (3) | 970 (97) | 6.5 (61) | 5 | 4 | 20.7 (30) | 2 |
|  | **120** | 28 (3) | 972 (97) | 6.1 (57) | 5 | 5 | 19.2 (28) | 2 |
|  | **150** | 25 (2) | 975 (98) | 5.8 (54) | 4 | 5 | 18.5 (27) | 2 |

### Colorectal cancers classified as FIT negative

Table 6 details cancers diagnosed following a FIT result under the current DG30 threshold of 10μg/g. Nine cancers (7.8%, 9/115) were diagnosed within six months of a negative FIT (all within 3 months) Five of these followed a change in bowel habit, four patients had anaemia, two had unexpected weight loss, and two had abdominal pain. Seven of these cancers were located in the rectum or sigmoid colon, and three presented with obstruction.

**Table 6:** Clinical characteristics of FIT negative cancers <10ug/g. 2WW: two-week-wait referral; CT: computed tomography; CIBH: change in bowel habit; Abdo: abdominal; IDA: iron deficiency anaemia; NET: neuroendocrine tumour.

| Age | Sex | FIT request details | FIT (µg/g) | Delay to lab (days) | FIT to diagnosis (days) | Diagnosis |
| --- | --- | --- | --- | --- | --- | --- |
| 69 | F | Change of bowel habit | 3.1 | <1 | 6 | 2WW, stenosing rectosigmoid |
| 64 | F | New onset diarrhoea and weight loss | 0.7 | <1 | 10 | 2WW, 20mm low rectal |
| 69 | M | Microcytic anaemia | 0.1 | 4-5 | 19 | 2WW, rectal |
| 83 | F | Irregular bowel habit | 0.8 | 3-4 | 23 | Obstruction, right hemicolectomy |
| 82 | M | Wind and abdominal pain | 3.8 | <1 | 24 | Obstruction, metastatic to liver |
| 32 | M | Weight loss, iron deficiency anaemia | 8.7 | 3-4 | 26 | 2WW |
| 82 | M | Anaemia | 0 | 3-4 | 28 | 2WW, sigmoid tumour on CT |
| 63 | M | Change bowel habit | 0.1 | 3-4 | 54 | 2WW, rectal tumour on CT |
| 74 | F | Hypertension, change in bowel habit | 1.5 | <1 | 91 | 2WW, rectal tumour |
| 53 | M | Abdominal pain | 0.1 | <1 | 188 | 2WW, terminal ileum NET |
| 48 | F | IDA | 1.7 | <1 | 374 | Obstruction, sigmoid tumour |
| 79 | F | ?PR bleed | 0.3 | <1 | 407 | 2WW, right hemicolectomy |

## Discussion

We report a large real-world diagnostic accuracy study including patients investigated with FIT for clinical features of colorectal disease in adult patients presenting to primary care in England. The sensitivity and specificity of FIT were both 91% for colorectal cancer using a FIT cut-off of 10ug/g, the threshold currently recommended by NICE. If investigated further, one in ten people with a positive FIT would be diagnosed with colorectal cancer whereas one person with cancer would not be referred for further investigation for every one thousand people tested. Selecting a higher threshold would mean that fewer patients without cancer would be eligible for referral for every person with cancer referred but a greater proportion of people not referred for further investigation would have cancer. For serious colorectal disease, the specificity and PPV of FIT were higher and the sensitivity and NPV were lower than for colorectal cancer.

### Strengths and Limitations

We present a large retrospective cohort study of FIT testing of English primary care patients to guide referral for colorectal cancer in the context of NICE DG30 guidelines. We ensured the population studied was relevant to DG30 by only including FIT requests originating in primary care. We have assumed that GPs have followed the guidance communicated to them about using FIT to triage “low risk” symptomatic patients. Although “high-risk” symptoms qualifying for urgent colonoscopy were noted in the clinical details, such as weight loss or anaemia, it can be assumed that GPs assessed these cases to be lower risk and not to qualify for fast-track referral, perhaps due to age, and that GPs required additional information to guide their management. This is consistent with previous literature recognising that symptoms and signs of disease form a “symptom continuum” and that interpretation of alarm symptoms vary between GPs (12). It is likely that the population included in our analysis accurately reflects patients selected for FIT testing in primary care in the context of the NICE DG30 guidance and it likely that the distinction between “high-risk” and “low-risk” symptoms is arbitrary when based on the symptom alone without taking into consideration characteristics such as duration or severity. As we were reliant on the limited clinical information provided with the test request, we were unable to investigate this further. Our results are consistent with previous reports that FIT more effectively categorises all symptomatic patients into ‘high risk’ and ‘low risk’ groups (17).

As this was a retrospective cohort study using electronic hospital records data from routine clinical practice, the majority of patients did not have a gold standard investigation at the same time as the FIT test to confirm whether serious colorectal disease was present or absent. All patients were followed up for evidence of subsequent pathology in hospital clinical, laboratory, radiology, endoscopy and pathology database between six and to 36 months after initial testing - a period that would reasonably allow significant pathology to be identified. Previous studies have used a shorter minimum follow-up period of three months and found similar results (12). We could have furthered out analysis by linking the dataset to the Public Health England’s National Cancer Registration and Analysis Service (NCRAS). NCRAS use multiple data sources to compile an accurate description of each cancer reported in England. Although we did not do this, by linking multiple local data sources for patients tested in a single central laboratory with a clearly defined geographical catchment area we increased the likelihood that serious disease diagnosed during the study period was captured.

Specimen preparation is a critical step in the overall performance of the FIT method (18,19). The methodological approach used in the present study involved collection of stool by the patient into a standard stool sample pot which was then sampled into the FIT method collection device by trained staff in the laboratory. This approach was adopted due to local concerns that patients may incorrectly use the collection device: even sampling by laboratory staff shows high imprecision (7, 19), and patient collection into the device precludes concurrent testing for other faecal tests, such as calprotectin. It is possible that, as a consequence of the delay between collection and arrival in the laboratory, some degree of degradation may have occurred and the quantitative FIT value may represent an underestimation of the true Hb concentration (20). However, the excellent discriminatory value of FIT (AUC of 0.94) and the low number of false negatives reported in this large real-world cohort indicates that Hb degradation is not a major limitation of clinical performance when using a low threshold of FIT (19).

### Comparison with existing literature

A number of recent studies have reported the diagnostic accuracy of FIT in symptomatic patients. A study including 3462 FIT samples from patients aged ≥30 years investigated in primary care for “low-risk” or “non-alarm” symptoms including change in bowel habit, abdominal pain, unexplained anaemia, and non-specific symptoms of fatigue and weight loss (12). 540 (15.6%) were positive at a threshold of 10µg/g but sensitivity and specificity were not reported. The PPV for colorectal cancer was 9.4% (95% CI: 7.0%-11.9%) overall, 6.05% (3.2%-8.7%) in women, and 13.3% (9.1%-17.5%) in men over 3-months of follow-up. The PPV in our study was similar 10.1% (8.2%-12.0%), 7.64 (5.24-10.0) in women and 12.5 (9.52-15.5) in men, despite the longer follow-up period. The same study reported a PPV for serious colorectal disease (colorectal cancer, high risk adenoma, and inflammatory bowel disease) of 14.7% (10.6%-18.9%) overall and in women 12.2% (8.1-16.2) and men 13.5% (10.6%-16.4%). The PPV for serious colorectal disease was higher in our study but remained lower for women than for men.

Others have reported the diagnostic accuracy of FIT in patients referred for colorectal investigation. A study from Scotland reported a sensitivity for colorectal cancer of 89.3%, specificity 79.1%, PPV 14.2%, and an NPV of 99.5% and for serious colorectal disease sensitivity 68.6%, specificity 83.6%, PPV 39.8%, and an NPV of 94.4% (11). In a Dutch cross-sectional diagnostic accuracy study (21), 810 patients referred for colonoscopy for suspicion of significant colorectal disease were investigated with point-of-care FIT using a haemoglobin threshold of >6µg/g. The sensitivity for significant colorectal disease (colorectal cancer, inflammatory bowel disease, diverticulitis, or advanced adenoma > 1 cm) was 67%; specificity 84%; PPV 47%; and NPV 92%. In a further retrospective English study of 430 non-consecutive patients (22) referred for urgent lower gastrointestinal investigation, the sensitivity for colorectal cancer was 84% and specificity 93%. The difference in performance of FIT found in our study is most likely to be due to our study including a lower risk spectrum of unselected symptomatic primary care patients - the population that NICE intended GPs to use

### Implications for research and practice

FIT could be used to further simplify referral pathways for people with suspected colorectal cancer. It could also provide a means to reprioritise patients with clinical features of colorectal disease who have had definitive investigation delayed, for example those currently awaiting colonoscopy deferred due to the COVID-19 pandemic. A recent English audit from Nottingham reported that a FIT ≥150ug/g triggered an immediate patient contact to arrange rapid investigation more expedient than urgent two-week-wait investigation (17). Based on our cohort, using a threshold of 150ug/g would lead to 3% of patients FIT tested being rapidly investigated to detect 54% of all undiagnosed colorectal cancer. Reducing the threshold to 120ug/g, the level used by the UK’s Bowel Cancer Screening Programme (BCSP) would lead to a similar percentage of patients being referred to detect an additional 3% of the underlying cancers. By setting the threshold at 10ug/g, 10% of the tested population would be further investigated to detect 91% of cancers. Furthermore, the PPV of 10% associated with a FIT of 10ug/g far exceeds the threshold of 3% above which NICE recommend further investigation for cancer in symptomatic patients, and the 8% threshold used in the BCSP. Therefore, symptomatic patients with positive FIT at a low cut off should be prioritised similarly or even ahead of asymptomatic patients with a positive FIT diagnosed as part of bowel cancer screening (23).

Use of FIT testing in colorectal low risk symptom pathways is likely to be controversial as was the introduction of gFOBT by NICE in 2015 (2, 3). However, patients and bowel cancer charities may be reassured by the improved performance of FIT compared to gFOBTs and with appropriate safety netting allow more rapid definitive testing for the at risk population (24). Commissioners of colorectal cancer services are likely to support this more efficient use of expensive endoscopic resource to target those at highest risk and to potentially reach underserved populations given the simplicity of the test (25). There will be large backlogs post COVID 19 and FIT use might ensure potential cancers are given higher priority as Trust and endoscopy services review deferred cases. Nevertheless labs may find a very substantial increase in FIT workload and will need to consider the logistics of getting FIT kits out to patients and back ideally via post in the COVID-19 pandemic, including making sure the correct clinician in primary or secondary care receives the result. There is likely to be comment by National professional bodies such as the Joint Advisory Group for Endoscopy (JAG), British Society of Gastroenterology, Association of Coloproctoloigists of Great Britain and Ireland on the use of FIT in this way: many societies have been supportive and again recognise that previous ways of using endoscopy services will not be sustainable in the post-COVID environment. Better to target high quality endoscopy at selected cases than try to carry out large volumes of lower quality colonoscopy. Finally a FIT threshold of 10ug/g is likely to be welcomed by endoscopists: it will clearly lead to a greater proportion of colonoscopies identifying important pathology or being therapeutic.

Previous research has considered the added value of interpreting FIT in combination with other clinical information, including additional faecal and blood tests. Combining negative FIT with a negative faecal calprotectin or a negative FIT and a normal haemoglobin were reported as safe approaches to ruling-out serious colorectal disease (11, 26). A multivariable model called COLONPREDICT was developed in Spain to predict colorectal cancer in patients referred for colonoscopy (27). It included twelve covariates derived from age, gender, FIT, blood haemoglobin, carcinoembryonic antigen, acetylsalicylic acid treatment, previous colonoscopy, rectal mass, benign anorectal lesion, rectal bleeding, and change in bowel habit). The Scottish FAST score is more simple, combining FIT, age and sex as a single test result which might indicate individual risk of colorectal cancer or serious colorectal disease (28). Despite their relative complexity compared to using FIT alone, the AUC for colorectal cancer was 0.92 (0.90-0.94) for COLONPREDICT and 0.87 (0.85–0.89) for FAST (29). Both of these scores show greater discriminative ability for colorectal cancer than the referral criteria included in the NICE NG12 suspected cancer guidelines (0.53 (0.49–0.57) (29). These studies have led researchers to the conclusion that referral criteria based on FIT measurement, either as a single test or within prediction models, are more accurate than symptom-based referral criteria (29). Based on the AUC of FIT alone (0.94 (0.91–0.97)) derived from our cohort of 9,896 English primary care patients tested with FIT for clinical features of colorectal cancer prior to referral, we consider FIT alone to perform with greater discrimination than using more complex approaches. We will investigate further but it allows a simple recommendation for primary care commissioners and clinicians to be made: a FIT result ≥10ug/g in a patient tested in primary care for suspected colorectal cancer or serious colorectal disease should prompt urgent referral for definitive colorectal investigation.

The false negative rate in this study was low at one in a thousand patients tested. Patients with a negative FIT, and persistent, new, or worsening symptoms should be encouraged to re-attend primary care within a defined period of time (e.g. within 4 weeks) so they are not lost to follow-up. By safety-netting in this way, clinicians may reconsider definitive colorectal investigation, investigation for other serious causes of abdominal symptoms, seeking specialist advice and guidance where necessary.

## Conclusion

FIT offers an appropriate triage test for use in primary care to investigate patients with symptoms of serious colorectal disease. A FIT threshold of 10ug/g would lead to 10% of patients tested being recommended for definitive investigation to detect 91% of the underlying cancers. Simple safety netting advice can guard against very infrequent false negative FIT results for colorectal cancer. FIT could be used to reduce pressure on urgent referral pathways by identifying patients who do not require further investigation for colorectal cancer, thereby controlling colonoscopy demand and reducing costs. FIT could be used in this way to reprioritise patients with lower colorectal cancer symptoms whose tests have been delayed by the COVID-19 pandemic.

## Data Availability

All data are available through Dr Brian Shine.

## Contributorship Statement

BDN, TJ, JO, JEE and BS conceived the study. TJ, BS, MP and SJ collected the data. BDN, JO, BS and TJ analysed the data. BDN prepared the first draft of the manuscript. All authors provided critical revisions to the manuscript.

## Ethical approval

This study was conducted as a service evaluation with registration, review and approval process within the OUH Datix governance structure (Service evaluation registration identifier: CSS-BIO-3-4730)

## Acknowledgements

We acknowledge Oxfordshire Clinical Commisioning Group (CCG).

